# PREOPERATIVE SERUM ALBUMIN LEVEL AS A PREDICTOR OF MORTALITY AND MORBIDITY AFTER VALVE REPLACEMENT SURGERY

**DOI:** 10.1101/2020.10.13.20206409

**Authors:** Md. Noor-E-Elahi Mozumder, Md. Mostafizur Rahman, Md. Rezwanul Hoque, Muhammad Nasif Imtiaz, Abu Jafar Md. Tareq Morshed, Md. Zanzibul Tareq, Md. Nahedul Morshed

## Abstract

**Background:** Serum albumin has a close correlation with degree of malnutrition which is associated with poor outcome and quality of life after cardiac surgery. Hypoalbuminemia is associated with increased wound infection, prolonged hospital stay and death after major surgery. Hence, preoperative serum albumin level can be utilized to upgrade risk models which will further benefit the cardiac surgical patients without extra financial burden. Objective of this study was to evaluate the role of serum albumin as a predictor of morbidity and mortality after valve replacement surgery.

**Methods:** This comparative cross-sectional study was carried out at the department of cardiac surgery in BSMMU. The study population was 50, with two groups having 25 patients each. Grouping of patients were done with respect to a preset cut off value for serum albumin. The period of study was from August, 2018 to February, 2020 and purposive sampling method was applied for this study. Data was collected by using a standardized semi-structured questionnaire and face to face interview.

**Results:** By demographic characteristics, mean age was significantly higher in group B (49.96±8.69 years) than in group A (41.60±11.16 years) (p=0.005). Mean BMI was lower in group B (20.88±3.71 kg/m^2^) than in group A (22.26±1.67 kg/m^2^), which was found statistically significant (p=0.006). In terms of postoperative outcome, total chest drain collection was significantly higher in group B (968.80±183.49 ml) than in group A (816.00±113.40 ml), (p=0.001). Duration of ICU stay were significantly longer in group B (4.60±0.76 days) than in group A (3.92±0.86 days) (p=0.005). Similarly, duration of hospital stay was significantly longer in group B (9.88±1.56 days) than in group A (8.64±0.81 days) (p=0.001). Overall morbidity was significantly higher in group B (48%) than in group A (20%) (p<0.05). Mortality rate was higher in group B (12%) than in group A (4%), but that was not found statistically significant (p>0.05). Pearson co-efficient correlation test showed strong inverse relationship of serum albumin with total chest drain, ICU stay and hospital stay following valve replacement surgery (r= −0.473, r= −0.448 & r= −0.487 respectively), which was most significant than age and BMI (p≤0.001). Multivariate logistic regression analysis was done to assess the predictive value of serum albumin level, age and BMI, where preoperative serum albumin level was found to be the most valuable predictor of postoperative morbidity after valve replacement surgery (B= −2.251, OR 0.105, 95% CI 0.011-0.986, p<0.05).

**Conclusion:** This study demonstrated that preoperative low serum albumin level is associated with increased morbidity and mortality after valve replacement surgery. Hence, preoperative serum albumin level can be used as a reliable predictor of postoperative outcome following valve replacement surgery.

## Introduction

Serum albumin is a commonly used indicator of nutritional status. It is a reliable and reproducible predictor of general surgical risk and has a close correlation with the degree of malnutrition. Malnutrition is a common problem in surgical patients that adversely affects outcome. It is strongly associated with postoperative complications and is a predictor of outcome, longer hospital stay and death ^[1]^. Preoperative malnutrition is more common in patients of valvular heart disease and is associated with increased morbidity and a poor quality of life after cardiac surgery. Therefore, it is important for operative risk stratification of patients with malnutrition, who may benefit from optimization of nutrition prior to surgery [2].

Albumin has been considered a negative acute-phase protein because its concentration decreases during injury and sepsis. Research on adult patients in various situations has shown the usefulness of this protein as a predictor of mortality ^[3]^. It is an important protein of human plasma. It represents 60% of the total plasma protein pool. Among the important physiological functions of albumin are maintenance of plasma oncotic pressure and the binding of several molecules, including fatty acids, bilirubin, metals, hormones, and drugs. It also acts as a free radical scavenger and reservoir for nitric oxide ^[4]^.

Recently it has been demonstrated that immediate postoperative hypoalbuminemia predicts poorer outcome and increased hospital stay after adult cardiac surgery. Postoperative hypoalbuminemia is common in patients undergoing major surgery. Several factors are responsible for this such as dilution secondary to fluid resuscitation, increased catabolism and reduction of synthesis, loss because of hemorrhage, gut loss, and redistribution secondary to altered vascular permeability. The degree of capillary permeability may be proportionate to the systemic inflammatory response, hence creates a greater rate of vascular permeability. In addition, increased blood loss and intraoperative administration of large fluid volumes further decrease serum albumin concentration. Therefore, patients having low serum albumin preoperatively are at greater risk of complications after surgery ^[5]^.

Hypoalbuminemia is associated with poor tissue healing, decreased collagen synthesis and granuloma formation in surgical wounds. These factors contribute to delayed wound healing, increased wound dead space, and create an environment that predispose to infection. It is also associated with impairment of macrophage activation and induces macrophage apoptosis, thus causes impairment of the innate immune response. Furthermore, it causes tissue edema due to reduction in plasma colloid osmotic pressure, which provides a medium for bacterial propagation. Combined, all these factors could promote wound infection after surgery and increase duration of stay in hospital ^[6]^.

Hypoalbuminemia before major non-cardiac surgery has been associated with increased wound infection, mortality and length of stay in hospital. It has been considered strong predictor of postoperative renal dysfunction and infection. Among body mass index and serum cholesterol as possible confounders, hypoalbuminemia has been shown to be an independent risk factor for poor short term surgical outcome. That’s why it is part of the commonly used prognostic system ‘APACHE’ (Acute Physiology and Chronic Health Evaluation), which predicts mortality among critically ill medical and surgical patients. Along with diabetes mellitus, low postoperative cardiac output, duration of operation, age, obesity and previous cardiac surgery, hypoalbuminemia indicates a high risk of infection among patients who undergo open-heart surgery ^[7]^.

Previously no study has been done in Bangladesh to see the association of serum albumin with postoperative morbidity and mortality after cardiac surgery. The aim of present study was to establish preoperative serum albumin concentration as a reliable predictor of morbidity and mortality after valve replacement surgery.

## Objective

To evaluate the role of preoperative serum albumin level as a predictor of postoperative morbidity and mortality following valve replacement surgery.

## Materials & Methods

This comparative cross-sectional study was carried out at the department of cardiac surgery in BSMMU. The study population was 50, with two groups having 25 patients each. Grouping of patients were done with respect to a preset cut off value for serum albumin (Group A: preoperative serum albumin level ≥ 3.5 gm/dl, Group B: preoperative serum albumin level < 3.5 gm/dl). The period of study was from August, 2018 to February, 2020 and purposive sampling method was applied for this study. Data was collected by using a standardized semi-structured questionnaire and face to face interview.

### Data Collection

All patients admitted in cardiac surgery department for elective cardiac valve replacement surgery without exclusion criteria were considered for study population. Patient who fulfilled the inclusion criteria and willing to enroll in the study was included in the study after receiving the proper consent. Preoperative serum albumin level was measured in each subject in Biochemistry laboratory, Bangabandhu Sheikh Mujib Medical University. Preoperative serum albumin level was measured according to Doumas, Watson and Biggs Method. Amount of specimen needed from each subject is 2 µL. To obtain this, after all aseptic precaution, venous blood sample was collected from each study subject in a disposable plastic syringe and immediately transferred to a sterile tube without any anticoagulant. Blood is allowed to clot freely in the tube. Then tubes were labeled with patients name, ID number & collection date and sent to Biochemistry laboratory immediately for centrifugation & further procedure. Patients who had preoperative serum albumin level ≥ 3.5 gm/dl were included in group A and who had preoperative serum albumin level <3.5 gm/dl.

### Data Analysis

Statistical analysis was conducted using Statistical Package for Social Science (SPSS) version 23.0 for windows software. Comparisons between groups were made with descriptive statistics like mean, frequency, range, standard deviation etc. and inferential statistics like Student’s t-test, Chi-Square test, Fisher’s exact test etc. Observations were recorded as statistically significant if a p-value is ≤0.05.

## Results

Total fifty (50) patients who underwent valve replacement surgery were evaluated in this study as per the inclusion and exclusion criteria. Venous blood samples to determine serum albumin level was collected preoperatively.

Among the study population mean age in group A was 41.60±11.16 years and in group B was 49.96±8.69 years. The difference in age between two groups was statistically significant (p<0.05). There was no statistical significance of gender between the two study groups (p>0.05). The mean BMI in group A was 22.26±1.67 kg/m^2^ and that in group B was 20.88±1.69 kg/m^2^. The findings were statistically significant (p<0.05).

Among the study population, total chest drain collection in group A and B were 816.00±113.40 ml and 968.80±183.49 ml respectively, which was statistically significant (p=0.001). Duration of ICU stay following surgery was longer in group B patients (4.60±0.76 days) compared to group A patients (3.92±0.86 days), which was statistically significant (p=0.005). Difference between the duration of hospital stay of the two groups was also statistically significant (p=0.001) as hospital stay in group B patients was longer (9.88±1.56 days) than group A patients (8.64±0.81 days).

Mortality rate was higher in group B (12%) in comparison to group A (4%), but this difference was statistically not significant (p>0.05). But there was statistical significance (p<0.05) in terms of morbidity in between two groups, where morbidity was higher in group B (48%) than group A (20%).

Pearson co-efficient correlation test for preoperative predictors and postoperative outcome showed strong inverse relationship of preoperative serum albumin level with total chest drain, ICU stay and hospital stay (r= −0.473, r=−0.448 & r= −0.487 respectively). It also showed preoperative serum albumin has most significant relationship (p<0.01) with total chest drain, ICU stay, hospital stay than age and BMI.

The relationship between baseline predictors and postoperative morbidity following valve replacement surgery were assessed by the use of logistic regression analysis. Among the variables, the preoperative serum albumin level was found to be the most valuable predictor (B= −2.251, OR 0.105, 95% CI 0.011-0.986, p<0.05).

## Discussion

This study aimed to find the impact of preoperative plasma albumin level for predicting in-hospital morbidity and mortality after valve replacement surgery. Valve replacement surgery was performed maintaining standard operative protocol and postoperative care was given to the patients of both groups as per standard protocol.

The demographic variables of the participating patients were recorded and analyzed. The mean age for group A was 41.60±11.16 years and group B was 49.96±8.69 years respectively, the difference was statistically significant (p=0.005). The age range of the patients of this study was from 22 years to 67 years. This study observed that the incidence of low serum albumin level and postoperative morbidity were more common in old age. A similar study carried out by Wei *et al*., showed serum albumin level and relation to age was statistically significant (p=0.005) ^[3]^.

In group A, approximately half of the population were male 13 (52%) and half were female 12 (48%). In group B, male and female patients were 16 (64%) and 09 (36%) respectively. The distribution of gender between two groups were not statistically significant (p=0.390). Similar findings were published by Koertzen *et al*., in their research ^[8]^.

When average BMI was compared between the two groups, it was 22.26±1.67 kg/m^2^ in Group A and 20.88±3.71 kg/m^2^ in Group B. The difference was statistically significant (p=0.006). This study observed that low serum albumin level and low BMI are associated with increased postoperative morbidity and mortality, which correlates to the findings of Bhamidipati *et al*., in their study ^[1]^. Demographic and anthropometric data are listed in Table 1.

**Table 1:** Comparison of demographic and anthropometric characteristics:

| Attributes | Group |  | p value* |
| --- | --- | --- | --- |
|  | Group A (n=25) | Group B (n=25) |  |
| <sup>a</sup>Age (in years) |  |  |  |
| Mean ± SD | $41.60 \pm 11.16$ | $49.96 \pm 8.69$ | 0.005<sup>s</sup> |
| Range | 22-62 | 37-67 |  |
| <sup>b</sup>Sex |  |  |  |
| Male | 13 (52%) | 16 (64%) | 0.390<sup>ns</sup> |
| Female | 12 (48%) | 9 (36%) |  |
| <sup>a</sup>BMI (kg/m<sup>2</sup>) |  |  |  |
| Mean ± SD | $22.26 \pm 1.67$ | $20.88 \pm 1.69$ | 0.006<sup>s</sup> |

Figure in the parentheses indicate percentage.
Data were analyzed using-
<sup>a</sup>Student's t-test and was presented as mean ± SD;
<sup>b</sup>Chi-square test ( $\chi^2$ ) was used to measure the level of significance.
\* $p > 0.05$ was considered not to be significant.
n= number of subjects s= significant ns= not significant
BMI= Body Mass Index

**Table 2:** Comparison of postoperative outcome:

| Postoperative outcome | Group |  | p value* |
| --- | --- | --- | --- |
|  | Group A (n=25) | Group B (n=25) |  |
| Postoperative ventilation time (hours) | $6.78 \pm 1.26$ | $7.16 \pm 1.49$ | 0.335<sup>ns</sup> |
| Total chest drain collection (ml) | $816.00 \pm 113.40$ | $968.80 \pm 183.49$ | 0.001<sup>s</sup> |
| Duration of ICU stay (days) | $3.92 \pm 0.86$ | $4.60 \pm 0.76$ | 0.005<sup>s</sup> |
| Duration of hospital stay (days) | $8.64 \pm 0.81$ | $9.88 \pm 1.56$ | 0.001<sup>s</sup> |

Data were analyzed using
Student's t-test and was presented as mean ± SD.
\* $p > 0.05$ was considered not to be significant.
n= number of subjects s= significant ns= not significant

**Table 3:** Comparison of postoperative morbidity and mortality:

| Attributes | Group |  | p value * |
| --- | --- | --- | --- |
|  | Group A (n=25) | Group B (n=25) |  |
| <sup>a</sup>Morbidity |  |  |  |
| Yes | 5 (20%) | 12 (48%) | 0.037<sup>s</sup> |
| No | 20 (80%) | 13 (52%) |  |
| <sup>b</sup>Mortality |  |  |  |
| Yes | 1 (4%) | 3 (12%) | 0.609<sup>ns</sup> |
| No | 24 (96%) | 22 (88%) |  |

Figure in the parentheses indicate percentage.
Data were analyzed using-
<sup>a</sup>Chi-square test ( $\chi^2$ ) & <sup>b</sup>Fisher's exact test and these were used to measure the level of significance
\* $p>0.05$ was considered not to be significant.
n= number of subjects s= significant ns= not significant

**Table 4:** Pearson correlation test for preoperative serum albumin, age, BMI, total chest drain, ICU stay and hospital stay:

| Attributes |  | Albumin | Age | BMI | Total drain | ICU stay | Hospital stay |
| --- | --- | --- | --- | --- | --- | --- | --- |
| Albumin | Pearson Correlation | 1 | -.338* | .425** | -.473** | -.448** | -.487** |
|  | p value |  | .016 | .002 | .001 | .001 | .000 |
|  | N | 50 | 50 | 50 | 50 | 50 | 50 |
| Age | Pearson Correlation | -.338* | 1 | -.111 | .210 | .305* | .308* |
|  | p value | .016 |  | .444 | .143 | .031 | .029 |
|  | N | 50 | 50 | 50 | 50 | 50 | 50 |
| BMI | Pearson Correlation | .425** | -.111 | 1 | -.069 | -.171 | -.287* |
|  | p value | .002 | .444 |  | .634 | .234 | .043 |
|  | N | 50 | 50 | 50 | 50 | 50 | 50 |
| Total drain | Pearson Correlation | -.473** | .210 | -.069 | 1 | .545** | .514** |
|  | p value | .001 | .143 | .634 |  | .000 | .000 |
|  | N | 50 | 50 | 50 | 50 | 50 | 50 |
| ICU stay | Pearson Correlation | -.448** | .305* | -.171 | .545** | 1 | .819** |
|  | p value | .001 | .031 | .234 | .000 |  | .000 |
|  | N | 50 | 50 | 50 | 50 | 50 | 50 |
| Hospital stay | Pearson Correlation | -.487** | .308* | -.287* | .514** | .819** | 1 |
|  | p value | .000 | .029 | .043 | .000 | .000 |  |
|  | N | 50 | 50 | 50 | 50 | 50 | 50 |

\* Correlation is significant at the 0.05 level (2-tailed).
\*\* Correlation is significant at the 0.01 level (2-tailed).

**Table 5:** Logistic regression analysis of predictors of postoperative morbidity following valve replacement surgery:

| Predictors | B | P value<sup>*</sup> | Odds ratio | 95% CI for odds ratio |  |
| --- | --- | --- | --- | --- | --- |
|  |  |  |  | Lower | Upper |
| Preoperative serum albumin level | -2.251 | 0.049<sup>s</sup> | 0.105 | 0.011 | 0.986 |
| BMI | 0.007 | 0.973<sup>ns</sup> | 1.116 | 0.681 | 1.489 |
| Age | 0.015 | 0.664<sup>ns</sup> | 0.734 | 0.949 | 1.086 |

<sup>\*</sup> $p>0.05$ was considered not to be significant.
s= significant
ns= not significant

Among the preoperative echocardiographic parameter ejection fraction (EF) was compared between group A and group B. The difference of EF in between the two groups was not statistically significant (p=0.945). Group A had a mean EF of 47.00±6.47 % and that of group B was 47.12±5.64 %. Cruz *et al*., reported similar findings in their study ^[7]^.

Regarding peroperative variables, mean cross clamp time, total CPB time & duration of surgery in group A were 76.44±33.91 minutes, 128.56±52.15 minutes and 249.00±73.78 minutes and in group B were 96.40±40.87 minutes, 154.80±62.22 minutes and 281.60±85.84 minutes respectively. Differences between two groups were statistically not significant (p=0.066, p=0.113, p=0.159 respectively). Engelman *et al*., published similar findings in their study ^[9]^.

Number of valve replaced and type of prosthetic valve used were also compared between two groups statistically. In most of the patients single valve replacement were done which is 19 patients (76%) in group A and 18 patients (72%) in group B whereas double valve replacement were done in 6 patients (24%) in group A and 7 patients (28%) in group B. The differences were statistically not significant (p=0.747). There were also no statistical significance in terms of whether mechanical or tissue valve were used as prosthetic valve between two groups (p=0.999). The findings corresponded with Wei *et al*., in their published reports ^[3]^.

Postoperative outcomes variables were also compared between two groups. Total chest drain collection were 816.00±113.40 ml in group A and 968.80±183.49 ml in group B, which was statistically significant (p=0.001). Mean duration of postoperative ventilator time were 6.78±1.26 hours in group A and 7.16±1.49 hours in group B, which was found statistically not significant (p=0.335). Engelman *et al*., published similar findings in their study ^[9]^.

In present study, statistical significance was found in terms of duration of ICU stay and duration of hospital stay between two groups. Duration of ICU stay were 3.92±0.86 days in group A and 4.60±0.76 days in group B, which was statistically significant (p=0.005). Duration of hospital stay were also statistically significant (p=0.001), where in group A it was 8.64±0.81 days and in group B was 9.88±1.56 days. Koertzen *et al*., found similar findings in terms of duration of ICU stay and duration of hospital stay with p value of p<0.001 & p<0.005 respectively ^[8]^.

In this study, postoperative complications like new onset arrhythmia (AF, VT and PVC), wound infection, stroke and re-exploration for bleeding were also studied to see any difference between two groups. Though wound infection was relatively higher in group B (16%) in comparison to group A (4%), there was no statistical significance (p=0.349). Other parameters were also statistically not significant (p>0.05). These findings corresponded with Bhamidipati *et al*., in their study ^[1]^.

Regarding overall morbidity and mortality between two groups, about 5 patients (20%) of group A and 12 patients (48%) of group B had developed one of the morbidities mentioned above, which was statistically significant (p=0.037). In this study, total 4 patients (8%) died out of 50 patients. Among them 1 patient (4%) was in group A and 3 (12%) were in group B. Though mortality rate was relatively higher in group B, but that was not statistically significant (p=0.609). Wei *et al*., found statistical significance in case of both morbidity (p=0.001) and higher mortality rate in hypoalbuminemic group (6.6% vs 3.1%, p=0.001) ^[3]^. As sample size was small in our study, mortality was not found statistically significant.

The Pearson co-efficient correlation test for preoperative serum albumin level, BMI, age, total chest drain, duration of ICU stay and duration of hospital stay showed significant inverse relationship of serum albumin level with total chest drain, ICU stay and hospital stay (r= −0.473, p=0.001; r= −0.448, p=0.001; r= −0.487, p<0.001 respectively). It also showed preoperative serum albumin level was most significant than BMI and age (significant at the 0.01 level) in relation to total chest drain, ICU stay and hospital stay. Bhamidipati *et al*., also found a strong inverse relationship between preoperative serum albumin level and postoperative outcome. They also found that albumin is a better predictor than BMI ^[1]^.

A logistic regression analysis was done to assess the predictive value of serum albumin level, BMI and age for postoperative morbidity following valve replacement surgery. Among the other variables serum albumin level was the most valuable predictor of postoperative morbidity following valve replacement surgery (OR 0.105, 95% CI, 0.011-0.986; p=0.049). Wei *et al*., provided similar information in their study (Wei *et al*., 2017). Rady *et al*., also found that preoperative serum albumin level was associated with postoperative morbidity after cardiac surgery in a multivariate analysis (OR 1.37, 95% CI, 1.04-1.81; p=0.03) ^[10]^.

As preoperative low serum albumin increases postoperative morbidity and mortality, further study should be done to assess the role of preoperative or peroperative albumin supplementation in patients with hypoalbuminemia in reducing postoperative complications.

## Conclusion

This study found preoperative serum albumin level as a reliable predictor of postoperative morbidity and mortality after valve replacement surgery. Thus preoperative serum albumin can be used as a screening tool for identifying individuals at added risk as well as will enable us to take necessary steps to reduce the risk of adverse events following valve replacement surgery.

## Limitations

a. This study was conducted upon a small sample size for a limited period of time.
b. This analysis was done in a single center of Bangladesh and the sample only represents a small fraction of patients undergoing cardiac surgery.
c. No follow-up was conducted after the discharge of the patient. So, adverse outcomes during follow-up period could not be assessed.

## Recommendations

I. Preoperative estimation of serum albumin level should be done routinely.
II. Serum albumin level should be corrected before surgery to reduce postoperative adverse events and it should be used as a screening tool to identify patients at added risk for adverse outcome after valve replacement surgery.
III. Large scale study with large sample size needed to validate the findings.
IV. We recommend preoperative serum albumin level as a reliable predictor of postoperative complications after valve replacement surgery and can be used as a risk assessment tool.

## Data Availability

Corresponding author has all data available with himself

