## Appendices for "PREOPERATIVE SERUM ALBUMIN LEVEL AS A PREDICTOR OF MORTALITY AND MORBIDITY AFTER VALVE REPLACEMENT SURGERY"

### APPENDIX A

### INFORMED CONSENT FORM

Objective:
The principle objective of this consent is to supply the patients all the necessary information to decide whether or not to participate in this study.

1. Name of study: "Preoperative serum albumin level as a predictor of mortality and morbidity after valve replacement surgery".

2. Name of investigator: Dr. Md. Noor-E-Elahi Mozumder

3. Place of study: Department of Cardiac Surgery, BSMMU.

4. Procedure:
   This study will be undertaken in department of Cardiac Surgery, BSMMU. This study will be carried out on patients who will have valvular heart diseases undergoing valve replacement surgery (VRS). There will be two groups- Group A will include patients having preoperative serum albumin level equal or more than 3.5gm/dl and Group B will include patients having preoperative serum albumin level less than 3.5gm/dl (2.5-3.4 gm/dl). Patients from these groups will undergo VRS and their postoperative complications (if any) will be observed. Surgery will be as per standard technique and will be carried out by same surgical team. The study duration will be of one and half year. Results of this study will predict outcome, possible complication and scope for better care and management for a large number of patients needing valve replacement surgery in future. If you agree to participate in this study, you will provide necessary information to the designated doctor. Results obtained from this study will benefit a large number of patients in near future.

5. Risk associated with study:
   There will be no deviation of care for patients undergoing this study. Preoperative investigation will be done in this hospital. But there is risk of complications related to anesthesia and surgery related.

6. Advantages of participation in the study:
   The patients will be directly benefited from this study as the patients will be treated in earliest possible way as per standardized procedure of care. This study will ensure better care for such category of patient in near future.

7. Alternative/compulsion in participation:
   Patient can withdraw himself/herself from the study at any time after participation. Patient can also decide regarding the matter of participation and will be allowed to make free decision without the influence of treating physician or hospital authority.

8. Cost:
   Patients will not have to bear any cost regarding this study.

9. Confidentiality:
   Information regarding patient's personal details and disease process will be kept confidential. All the information will be stored in departmental office and all the documents will be used in data analysis.

10. Participation in study:
    Participation in this study is voluntary. Patient may participate; withdraw from participation at any time during the study period. Even if the patient withdraws from the study, there will be no variation from standard technique. Signing this form will not violate any of the standard human rights.

11. Responsibility of researcher and medical care:
    If the patient gets sick or seriously ill while participating in the study, the researcher will bear all the expenses for the treatment.

12. Questionnaire:
    Regarding the treatment procedure or study, patient is allowed to ask any questions regarding the procedure or the study and researcher will try to clarify as much as possible.

13. Declaration of consent:
    I am concerned about my illness and different modalities of treatment. I am also aware of the surgery and its possible outcomes. I am also informed about the importance of study carrying on me. After knowing every pros and cons, I give consent in sound state of mind without any external influence. I give consent to the investigator Dr. Md. Noor-E-Elahi Mozumder with the opportunity or chance of withdrawal from this study at any stage.

____________________

Signature of Patient
Name:
Age:
Address:
Phone No.:

<u>Witness Name and Signature</u>:
1.
2.

____________________

Signature of Researcher
Name:

### APPENDIX B

#### অবগতিক্রমে সম্মতি পত্র

**উদ্দেশ্য:**

এই সম্মতি পত্রের উদ্দেশ্য হল আপনাকে প্রয়োজনীয় তথ্য প্রদান করা, যা আপনাকে সিদ্ধান্ত নিতে সাহায্য করবে যে আপনি এই গবেষণায় অংশগ্রহণ করবেন কি না।

১. গবেষণার নাম ঃ “প্রিঅপারেটিভ সেরাম এল্বুমিন লেভেল এস এ প্রিডিক্টর অফ মরটালিটি এন্ড মরবিডিটি আফটার ভালভ রিপ্লেসমেন্ট সার্জারি”।

**২. গবেষকের নাম ঃ ডাঃ মোঃ নুর-এ-এলাহি মজুমদার**

৩. **স্থান ঃ** কার্ডিয়াক সার্জারি বিভাগ, বঙ্গবন্ধু শেখ মুজিব মেডিকেল বিশ্ববিদ্যালয়

৪. **পদ্ধতি ঃ**

এই গবেষণাটি বঙ্গবন্ধু শেখ মুজিবুর মেডিক্যাল বিশ্ববিদ্যালয়ে কার্ডিয়াক সার্জারি বিভাগে, যে সব রোগী ‘ভালভুলার হার্ট ডিজিজ’ রোগ নিয়ে ভর্তি হবেন, তাদের উপর পরিচালিত হবে। এই গবেষণাটির মাধ্যমে এই রোগ, আরো উন্নত চিকিৎসা পদ্ধতি, অপারেশনের পরিণতি ও সম্ভাব্য জটিলতা অনুসন্ধান করা হবে এবং গবেষণার প্রাপ্ত তথ্য দেশের বিপুল সংখ্যক রোগীদের উন্নত চিকিৎসার কাজে লাগবে। রোগীদেরকে দুই গ্রুপ-এ ভাগ করা হবে। গ্রুপ- ‘এ’ যাদের সেরাম এল্বুমিন লেভেল ৩.৫ গ্রাম/লিটার-এর সমান বা বেশি । গ্রুপ-বি যাদের সেরাম এল্বুমিন লেভেল ৩.৫ গ্রাম/লিটার-এর কম । দুই গ্রুপের সার্জারী হবে আর রোগীদের পরিণতি গবেষণা হবে। আপনি যদি গবেষণায় অংশগ্রহণ করতে সম্মত থাকেন তাহলে গবেষণায় নিয়োজিত চিকিৎসককে তথ্য প্রদান করবেন। গবেষণায় প্রাপ্ত তথ্য দেশের বিপুল সংখ্যক রোগীদের উন্নত চিকিৎসার কাজে লাগবে।

**৫. গবেষণার ঝুঁকি ঃ**

এই গবেষণায় অংশগ্রহণকারীদের প্রচলিত ও নির্ধারিত চিকিৎসার কোন ব্যতিক্রম হবে না। অপারেশনের প্রস্তুতিমূলক পরীক্ষাসমূহ এই হাসপাতালে করা যাবে। এনেস্থেসিয়াজনিত জটিলতা এবং অপারেশন পরর্বতী ইনফেকশন জটিলতার ঝুঁকি রয়েছে।

**৬. গবেষণায় অংশগ্রহণের সুবিধাদি ঃ**

এই গবেষণায় অংশগ্রহণ করলে আপনি ব্যক্তিগতভাবে সরাসরি লাভবান হতে পারবেন। সঠিক ও উন্নত চিকিৎসার মাধ্যমে আপনি দ্রুত আরোগ্য লাভ করবেন। এই গবেষণা বাংলাদেশের চিকিৎসকদের এই রোগ ও তার চিকিৎসা সর্ম্পকে আরও জানতে সাহায্য করবে।

**৭. বিকল্প/ অংশগ্রহণের বাধ্যবাধকতা ঃ** এই গবেষণায় অংশগ্রহণ করা কিংবা না করার ব্যাপারে অথবা অংশগ্রহণ করার পর যে কোন সময়ে আপনি বা আপনার রোগীকে এই গবেষণা থেকে সরিয়ে নেয়ার ব্যাপারে আপনার পূর্ণ অধিকার থাকবে।

**৮. খরচ ঃ** এই গবেষণায় অংশগ্রহণ করার জন্য আপনার কোন খরচ নাই বা আপনাকে কোন অর্থ দেয়া হবে না।

**৯. গোপনীয়তা ঃ**

এই গবেষণা চলাকালীন বা পরর্বতীতে রোগীর সমস্ত তথ্য কঠোরভাবে গোপন রাখা হবে। পরর্বতী ফলোআপ বা অন্যান্য প্রক্রিয়ার জন্য আপনাকে নির্দিষ্ট তারিখ দেয়া হবে। আপনার সব তথ্য এই বিভাগের অফিসে সংরক্ষণ করা হবে। যা শুধুমাত্র এই গবেষণা ব্যতিত অন্য কোন কাজে ব্যবহার বা প্রকাশ করা হবে না।

**১০. গবেষণার অংশগ্রহণ ঃ**

এই গবেষণায় অংশগ্রহণ সম্পূর্ণ স্বেচ্ছামূলক। রোগী গবেষণায় অংশগ্রহণ করতে অস্বীকৃতি জানাতে পারেন অথবা গবেষণা চলাকালীন যে কোন সময়ে নিজেকে প্রত্যাহার করে নিতে পারেন। তাতে আপনার চিকিৎসার কোনরূপ তারতম্য হবে না। এই ফরমে স্বাক্ষর করলে আপনার আইনগত কোন অধিকার খর্ব হবে না।

**১১. গবেষণাকারীর দায়িত্ব ও মেডিকেল কেয়ার ঃ** এই গবেষণার অংশ গ্রহণের ফলে আপনি যদি আহত বা অসুস্থ হয়ে পড়েন, তা হলে গবেষণাকারী চিকিৎসার সকল ব্যয়ভার বহন করবেন।

**১২. প্রশ্নাবলী ঃ**

এই গবেষণা বা চিকিৎসা সর্ম্পকে যে কোন প্রশ্ন আপনি এই গবেষণার চিকিৎসককে জিজ্ঞাসা করতে পারবেন। যার উত্তর এই চিকিৎসক প্রদান করতে যথাসাধ্য চেষ্টা করবেন।

**১৩. সম্মতির স্বীকারোক্তি ঃ**

আমি আমার অসুস্থতা ও এর বিভিন্ন চিকিৎসা পদ্ধতি সর্ম্পকে বিস্তারিত অবগত হয়েছি। আমি সার্জারি ও এর সম্ভাব্য ফলাফল সর্ম্পকেও অবগত হয়েছি। আমাকে নিয়ে করা গবেষণা ও এর গুরুত্ব সর্ম্পকে আমাকে বিস্তারিত জানানো হয়েছে। আমি সবকিছু জানার পর স্বেচ্ছায়, স্বজ্ঞানে এবং বিনা প্ররোচনায় উক্ত অপারেশন এবং গবেষণা করার অনুমতি এই গবেষণার গবেষক **ডাঃ মোঃ নুর-এ-এলাহি মজুমদার** -কে দিচ্ছি। আমি যে কোন সময় উক্ত গবেষণা হতে নিজেকে প্রত্যাহার করার অধিকার সংরক্ষণ করি।

**রোগী/ অভিভাবকের স্বাক্ষর /বৃদ্ধাঙ্গুলির ছাপ**

নাম ............................................................ বয়স ..............

ঠিকানা ঃ ........................................................ ফোন নং ...................................

**সাক্ষীর স্বাক্ষর ও নাম** **গবেষণাকারীর স্বাক্ষর ও নাম**

১)

২)

### APPENDIX C

#### Data Collection Tool

#### Data Collection Sheet

#### QUESTIONNAIRE IN ENGLISH

**Title: Preoperative serum albumin level as a predictor of mortality and morbidity after valve replacement surgery.**

Name of Researcher: Dr. Md. Noor-E-Elahi Mozumder, Resident, Phase-B, MS (CVTS)

Name of Institute: Bangabandhu Sheikh Mujib Medical University (BSMMU), Dhaka.

**Case no:** ☐

**Section A: Information Regarding Patient Profile**:

**Name of the patient***:* ☐

**Age:** ☐ years **Sex:** Male=1, Female =2 ☐

**Address:** ☐

**Marital status:** 1. Married 2. Unmarried 3. Divorced 4. Widow 5. Other (Please specify) ☐

**Religion:** 1.Islam 2.Hindu 3.Buddhish 4. Christian 5. Other (Please specify ☐

**Occupation**: 1.Housewife 2.Service 3.Business 4.Farmer 5.Student 6. Other (Please specify) ☐

**Registration/ PIN no:** ☐

**Bed:** ☐ **Ward:** ☐ **Unit:** Orange=1, Red=2, Blue=3, Violet=4 ☐

**Contact/Mobile no:** ☐☐☐☐☐☐☐☐☐☐☐☐☐☐☐☐

**Date of Admission:** ☐☐☐☐☐☐☐☐

**Date of Procedure/Operation:** ☐☐☐☐☐☐☐☐

**Date of Data Collection:** ☐☐☐☐☐☐☐☐

**Date of Discharge:** ☐☐☐☐☐☐☐☐

**Section B: Information Regarding Clinical History:**

**Presenting Complaint/s:** [Yes=1, No=2]

Dyspnea: ☐ Duration: ☐

Palpitation: ☐ Duration: ☐

Chest pain: ☐ Duration: ☐

Syncope: ☐ Duration: ☐

Fatigue: ☐ Duration: ☐

**History of Past Illness:** [Yes=1, No=2]

Operation: ☐ Blood Transfusion: ☐

**Personal History:** [Yes=1, No=2]

Smoker: ☐ If yes- ☐ Pack/year

Alcoholic: ☐ Other: ☐

**Medical History:** [Yes=1, No=2]

Hypertension: ☐

DM: ☐

IHD/Stroke/MI: ☐

Heart Failure: ☐ Other (Please specify): ☐

**Drugs History:** [Yes=1, No=2]

Calcium channel blocker:

Angiotensin receptor blocker/ACEI:

Beta blocker:

Diuretics:

Anti-platelet:

**<u>Section C: Information Regarding Clinical examination and Findings</u>**:

Anaemia: Absent =1, Present =2 Jaundice: Absent=1 Present =2

Cyanosis: Absent=1, Present =2 Oedema: Absent=1, Present =2

JVP: Normal=1, Raised =2

Body Surface area: $m^2$

Body weight: kg Height: cm BMI: kg/ $m^2$

Pulse: beats per min Regular 1, Irregular =2

Blood pressure: mm of Hg

**Heart:** 1st Sound: Normal =1 Soft=2, Loud=3, absent=4

2nd sound: Normal =1, Soft=2, Loud=3, absent=4

Murmur: Absent =1, Systolic =2, Diastolic =3, Pan systolic =4

**Lungs:** Breath sound: Vesicular=1, Bronchial =2

Added Sound: Crepitation =1, Rhonchi=2

**<u>Section D: Information Regarding Preopearative Investigations:</u>**

Hb: gm/dl ESR: mm in 1st hour, CRP: mg/L

S. Creatinine: mg/dl FBS/RBS/2HABF: mmol/L

Serum Albumin: gm/dl Blood Group:

Lipid Profile: 1. Normal 2. Abnormal ☐ If abnormal, please specify: ☐

Liver Function Test: 1. Normal 2. Abnormal ☐ If abnormal, please specify: ☐

Thyroid Profile: 1. Normal 2. Abnormal ☐ If abnormal, please specify: ☐

Viral Markers: 1. Positive 2. Negative ☐ If positive, please specify: ☐

Chest X-ray P/A view: 1. Normal 2.Cardiomegaly ☐ Other (Please specify) ☐

ECG: 1. Normal 2. IHD 3. MI 4. LVH 5.RVH 6.AF 7.LBBB 8. RBBB

9. Hyperkalemia/Hypokalemia ☐ Other (Please specify) ☐

Echocardiogram (2D, M-mode & Color Doppler) findings:

☐

Coronary Angiogram:

**Diagnosis:** ☐

**<u>Section E: Information Regarding Procedure</u>:**

**Date of operation:** ☐☐☐☐☐☐☐☐

**Indication of surgery:** ☐

**Operative findings:** ☐

Duration of surgery: ☐ min

Cross clamp time: ☐ min

Cardiopulmonary bypass time: ☐ min

Peroperative arrhythmia (AF/VT/Arrest): 1. Yes 2. No [ ] (If yes, please specify) [ ]

Peroperative inotropes used: 1. Yes 2. No [ ]

**<u>Section F. Information Regarding Postoperative Condition:</u>**

Postoperative recovery: 1. Eventful 2. Uneventful [ ]

Duration of ventilation: [ ] hours

Postoperative vital parameters: 1. Normal 2.Abnormal [ ]

If 'Abnormal', please specify: [ ]

Postoperative laboratory profile: 1. Normal 2. Abnormal [ ]

If 'Abnormal', please specify: [ ]

Postoperative bleeding: [ ] ml /24 hours

Total chest drain collection: [ ] ml

Postoperative blood transfusion: [ ] unit(s)

Re-exploration done for hemodynamic instability: 1. Yes 2. No [ ]

Postoperative complications: 1.Yes 2. No

Excessive bleeding [ ] Thrombo-embolism [ ] Stroke [ ] Infection [ ]

MI [ ] AF [ ] VT [ ] Renal Failure [ ] Other (please specify) [ ]

Duration of ICU stay: [ ] Days

Duration of hospital stay: [ ] Days

Hospital Morbidity/Mortality: 1.Yes, 2.No [ ]

If yes, please specify: [ ]

**Signature of Investigator**

**Date:**

### APPENDIX D

#### CHECK LIST

**Title: Preoperative serum albumin level as a predictor of mortality and morbidity after valve replacement surgery.**

Name of Researcher: Dr. Md. Noor-E-Elahi Mozumder, Phase-B (CVTS)

Name of Institute: Bangabandhu Sheikh Mujib Medical University (BSMMU), Dhaka.

Case no.:

| Sl no. | Parameters/Investigations | Availability | | Value |
|---|---|---|---|---|
| | | Yes | No | |
| | Age | | | |
| | Sex | | | |
| | Weight in Kg | | | |
| | Height in cm | | | |
| | Hypertension | | | |
| | Diabetes Mellitus | | | |
| | History of smoking | | | |
| | Hemoglobin | | | |
| | Serum albumin | | | |
| | CRP | | | |
| | ALT | | | |
| | Serum bilirubin | | | |
| | Cross clamp time | | | |
| | CPB time | | | |
| | Duration of surgery | | | |
| | No. of valve replaced | | | |
| | Type of prosthetic valve used | | | |
| | Total chest drain | | | |
| | Wound infection | | | |
| | Postoperative arrhythmia | | | |
| | Duration of ICU stay | | | |
| | Duration of hospital stay | | | |
| | Morbidity | | | |
| | Mortality | | | |

### APPENDIX E

#### IRB Clearance Certificate

বঙ্গবন্ধু শেখ মুজিব মেডিক্যাল বিশ্ববিদ্যালয়
**Bangabandhu Sheikh Mujib Medical University**

রেজিস্ট্রার অফিস
Office of the Registrar

*NO. BSMMU/2019/333*

*Date:14-01-2019*

**Dr. Md. Noor-E-Elahi Mozumder**
MS (Cardiovascular and Thoracic Surgery) Phase-B, Resident
Department of Cardiac Surgery
Bangabandhu Sheikh Mujib Medical University
Shahbag, Dhaka-1000

Sub:**Institutional Review Board (I.R.B) Clearance.**

With reference to your application on the above mentioned subject, this is to inform you that your Research Proposal entitled **"Impact of preoperative serum albumin level as a predictor of mortality and morbidity after valve replacement surgery"** has been reviewed and approved by the Institutional Review Board (IRB) of Bangabandhu Sheikh Mujib Medical University in its 171th meeting held on 17 November 2018.

You are requested to follow the Institutional Review Board (IRB) guidelines.

**Expected Examination date July' 2020.**

14/1/19

**(Dr. Ferdous Alam)**
Member Secretary
Institutional Review Board
BSMMU, Shahbag, Dhaka

ব্লক বি (৩য় তলা), শাহবাগ, ঢাকা-১০০০, বাংলাদেশ, টেলিফোন ঃ ৮৬১২৫৫০-৪, ৮৬১৪৫৪৫-৯, ফ্যাক্স ঃ ৮৮০-২-৮৬২৪৮১৭,
Block B (2nd Floor), Shahbag, Dhaka-1000, Bangladesh, Tel : 8612550-4, 8614545-9, Fax : 880-2-8624817,
